# Social impact of the JACO wheelchair-mounted robotic arm on users and their caregivers

**DOI:** 10.1101/2024.09.18.24313906

**Authors:** Orthelo Léonel Gbètoho Atigossou, Julie Faieta, Maëlle Corcuff, Alexandre Campeau-Lecours, Véronique H. Flamand, François Routhier, Jason Bouffard

**Affiliations:** School of Rehabilitation Sciences, Faculty of Medicine, Université Laval, Quebec City, Canada; Center for Interdisciplinary Research in Rehabilitation and Social Integration (Cirris), Centre Intégré Universitaire de Santé et de Services Sociaux de la Capitale Nationale (CIUSSS-CN), Quebec City, Canada; Department of Rehabilitation Science and Technology, University of Pittsburgh, Pittsburgh, USA; Department of Kinesiology, Faculty of Medicine, Université Laval, Quebec City, Canada

**Keywords:** Wheelchair Mounted Robotic Arm, Robotics, JACO arm, Social impact, Upper limb disability

## Abstract

The efficacious implementation of robotic assistive technologies must be built on a thorough understanding of stakeholders’ experiences and perceptions. This study provides an in-depth insight into the experiences and perceptions of users of the JACO wheelchair-mounted robotic arm and those of their caregivers. A sample of JACO users (n = 21; Female : 6; Male : 15) and caregivers (n = 11; Female: 2; Male: 9) participated in individual interviews used to gain qualitative insight into the impact of JACO on their day-to-day lives. Interview transcripts were analyzed using a hybrid deductive-inductive coding process. Thematic analysis was conducted in accordance with the *Consortium on Assistive Technology Outcomes Research* (CATOR) taxonomy. This article exclusively reports data on the social impact of the JACO wheelchair-mounted robotic arm. In addition, participants completed three questionnaires to gather more objective data for quantitative assessment, these included the *Caregiver Assistive Technology Outcome Measure* (CATOM), a sociodemographic questionnaire, and a home-based questionnaire to assess the social impact of using JACO. Findings pointed to highly varied experiences among participants, including instances of positive, negative, and absence of effects from the use of JACO. Participants’ feedback fell within two broad categories, *Human Assistance,* and *Cost and Use of Resources*. This study provides nuanced and varied insight into the spectrum of the social impact of using JACO as perceived by users and their caregivers, highlighting the importance of considering each user as an individual with unique experiences and needs. Continued research is needed to assess the generalizability of these findings.

## Introduction

Caring for an individual with a chronic condition or physical limitation can be highly rewarding but it can also be time-consuming and physically and emotionally taxing. Informal or unpaid caregivers, often family members, have been found to experience negative health effects such as burden, depression, or anxiety [1, 2]. In addition to caregivers, assistive technologies can be used to support autonomy for those living with functional limitations [3]. Enhancing the autonomy of an individual with functional limitations can impact the amount of time and assistance required from an informal caregiver. In this manner, the use of an assistive technology device may indirectly influence the social participation, quality of life, and physical or mental health of informal caregivers potentially allowing them to return to their role as a family member or friend rather than a caregiver [4]. It is important to note the far-reaching effects of assistive technology device use, extending beyond the individual who uses the device. Jutai et al. highlighted that assistive technology outcomes affect both the user and those around him or her, including caregivers and the larger society [5]. The social impact can also include economic burdens (e.g., costs related to devices and services) and benefits (e.g., reduced need for human assistance and institutionalization) [5]. However, the effectiveness of an assistive device can vary over time due to changes in users’ needs, lifestyles, context, and preferences. If poorly suited to the individualized needs of the user, an assistive device could induce unwanted outcomes, such as added burden, frustration, and complexity. Therefore, effective technology evaluation is crucial for the successful implementation of assistive devices. In alignment with the *Human Activity Assistive Technology* (HAAT) model, assistive technology evaluation must consider the effectiveness of the assistive device for meeting the user’s needs in their day-to-day life activities and within the context of their individualized settings [6].

Upper limb disabilities have been linked to reduced autonomy and limited social participation in individuals affected by various neurological disorders [3] such as multiple sclerosis [7], spinal cord injury [8], and Duchenne’s muscular dystrophy [9]. Examples of currently available upper extremity assistive devices include reachers, orthoses, dynamic arm supports and robotic arms [3, 10]. Wheelchair-mounted robotic arms (WMRA), such as the JACO arm by *Kinova* (Canada) and the iArm by *Exact dynamics* (Netherlands), are a specific type of robotic devices that attach to a wheelchair and assist users in performing functional tasks that would otherwise be challenging or impossible due to limited upper extremity function. These tasks include picking up and grasping objects needed for feeding, self-care, work-related activities, or leisure tasks. In accordance with best practice in healthcare, evidence is needed to confirm the effectiveness of WMRA’s in the intended user populations. Unfortunately, to date, a limited number of studies have assessed WMRAs in real-world environments [10]. Existing studies show WMRA use in various routine activities by both novice and experienced users [11, 12]. They also provided insight on the potential impact of WMRA use on caregivers as perceived by the caregivers themselves [10], the users [10, 13], or by an independent observer [11]. Findings have indicated that WMRA use may lead to decreased need for human (*caregiver*) assistance for the care recipient (*user*). However, one of these studies indicated that informal caregivers (n=5) reported limited or no impact of a WMRA on their burden [10]. Because these studies yield conflicting results, – perhaps due to small sample sizes, differences in the assessment contexts (*Canada vs. European countries*), the outcome assessments used, or the perspectives assessed – additional research is needed to better understand the perceived value of WMRA for both users and caregivers.

Therefore, the present study sought to add to the limited body of available literature on stakeholders (*users and caregivers*) perceived value of WMRA use. This paper reports on a segment of a larger study that assessed outcome domains suggested in the taxonomy by Jutai et al. (*i.e., effectiveness, subjective-well-being and social significance*) [5]. The specific objective of this study was to document the social impact of an WMRA, as perceived by users and caregivers.

## Materials and methods

### Research design

An embedded mixed research design is employed to investigate the perceptions of users (*individual with functional limitations who utilizes a WMRA*) and caregivers of WMRA use in daily life and within the community context [14]. With this methodology, both quantitative and qualitative data are utilized. However, the qualitative perspective of WMRA users and their caregivers was the primary focus of interest given the exploratory nature of the study.

### WMRA technology and participants

The specific investigated WMRA was the JACO arm [15]. This WMRA interfaces with a variety of control hardware options to accommodate user preferences. These control options include joystick, head array and sip-and-puff options. A convenience sample was identified through a specific WMRA vendor (*Kinova*). All JACO users were older than 14 years of age (*as recorded by Kinova*) and living in the province of Quebec. Each user included in the study was asked to refer their primary informal (*unpaid, often a family member*) caregiver so that caregivers’ perspectives on the JACO arm could be investigated as well. Informal caregivers were included if they aided the user for at least 4 hours per week. All participants (users and caregivers) completed informed consent (ascent for minor participants) prior to participation in the study. The study was approved by the research ethics board of the *Centre intégré universitaire de santé et de services sociaux de la Capitale-Nationale*, (Quebec City, Canada; Ethics: ID# 2018-608). Once consented, participants completed a series of structured questionnaires as well as a semi-structured interview. The assessment tools of the larger study were selected based on Jutai et al. taxonomy [5] to assess all dimensions of assistive technology outcomes. Given the scope of the present paper, only assessment tools used to describe the study population and to assess social outcomes have been included.

### Samples description

A total of 31 JACO users within the province of Quebec were considered for recruitment, and 21 agreed to participate. Among the 10 users who did not participate, 1 refused, 1 showed interest, but did not ultimately participate, and 8 could not be reached (e.g., no reply and no available email address). Eleven caregivers (from 10 included users) were recruited via referral from their respective JACO users. Caregivers of the remaining users were either not interested in participating, unable to coordinate to participate in study assessments, or in some cases, users had no informal caregivers. As shown in Table 1, the sample was heterogeneous in terms of diagnosis, age, gender, time since onset of disability, and time since acquiring JACO. All caregivers were relatives of the users (primarily maternal) and were therefore older than users. Most caregivers live with the user and spend more than 30 hours of caregiving each week (n = 8 [73%]).

**Table 1.**
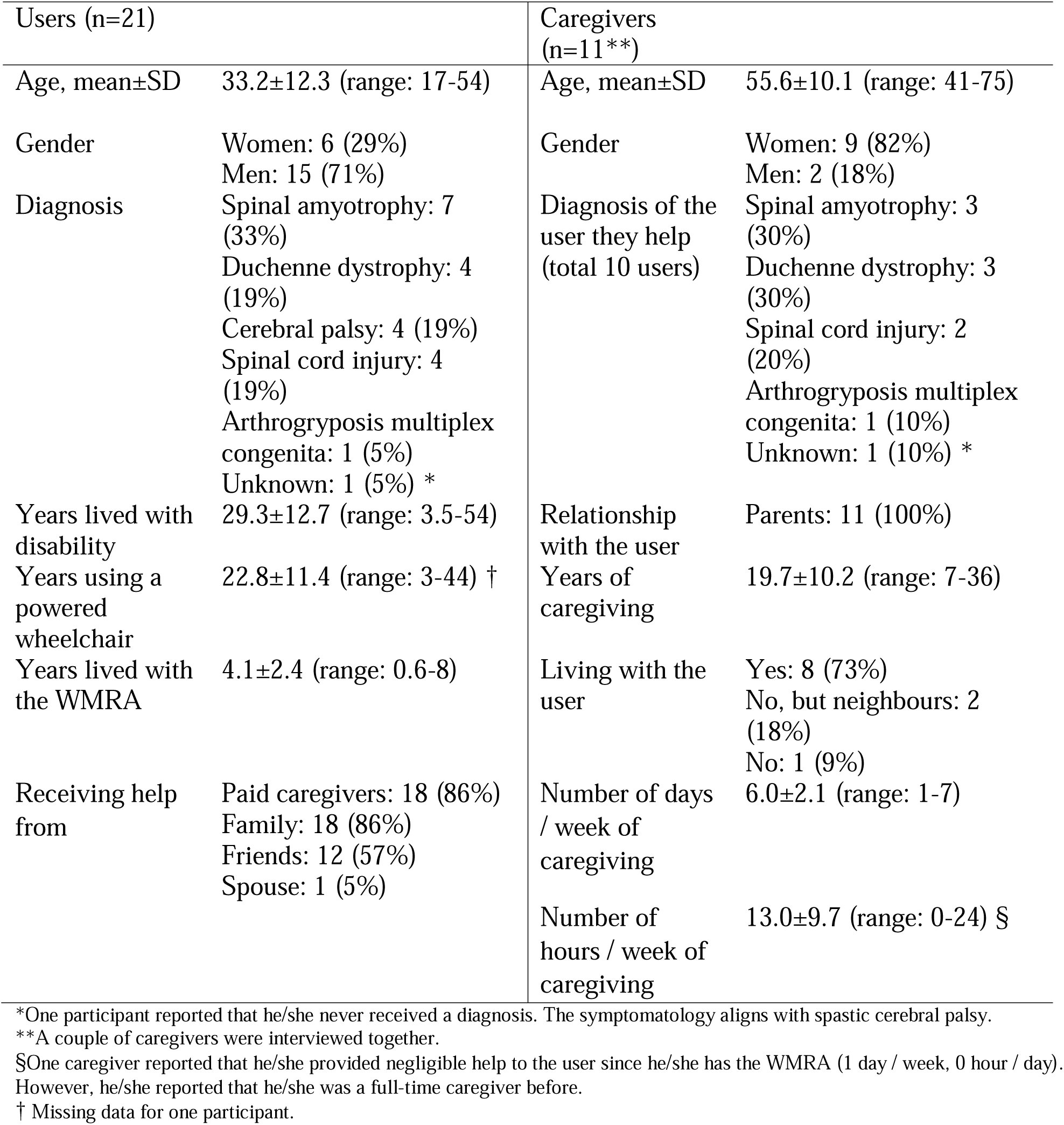
Samples description.

### Taxonomy of Assistive Technology Device Outcomes

Jutai et al. [5] proposed the *Consortium on Assistive Technology Outcomes Research* (CATOR) taxonomy that helps classify outcomes to promote consistency in how assistive technology devices affect the user, caregivers and the society as a whole. This taxonomy accommodates a wide variety of assistive technology device applications, encompassing various user populations, devices, services, and usage contexts [5]. Three main concepts are addressed: *effectiveness* (on functions, environmental factors and user longevity), *social significance* (impact on caregiving, costs, residential care placement, service utilization and device utilization), and *subjective well-being* (psychological functioning, quality of life and satisfaction).

### Individual Interviews

Prior to the study, group discussions and interviews were conducted with occupational therapists (n = 3), rehabilitation services managers (n = 2) as well as representatives from an industrial partner (n= 6) and from an assistive technology funding agency (n = 1) to determine the information they would seek to gather from JACO users and their caregivers. Interview guides were subsequently developed for users and caregivers based on stakeholders’ needs. The interview guides were compared to the taxonomic of Jutai et al. to ensure coverage of all assistive technology outcome domains [5]. The interview guides were not pre-tested before data collection to maximize the amount of data collected from this rare clinical population. However, we solicited feedback during the initial interviews with users and caregivers, which prompted minor modifications such as rewording and removing redundant questions. The resulting questions can be found in the supplementary materials. Both guides assess effectiveness, subjective well-being, and social significance outcomes related to JACO use from both users’ and caregivers’ perspectives [5]. However, more questions were devoted to the social significance impact for caregivers, given their unique perspective on the indirect effects of the JACO arm on their lives (note that guiding questions for caregivers and users varied). The interviews were conducted either in person or through videoconferencing software (*Zoom*).

### Structured questionnaires: User’s perspective

A series of questions assessing the sociodemographic profile (i.e., age, gender, occupation, residence), and clinical profile (diagnosis, time since onset of disability, starting to use a power wheelchair and the JACO arm) of the users were developed for this project based on previous studies on WMRAs [13, 16]. Users’ perceptions of the social significance of WMRA outcomes were collected via two user-completed questions assessing: 1) the impact of JACO on the caregivers’ perception of user safety when the user is alone, and 2) the user’s ability to remain in the community (rather than being institutionalized) [16].

### Structured questionnaires: Caregiver’s perspective

Sociodemographic information (i.e., age, gender, occupation) and caregiving-related details (i.e., relationship with the user, duration and frequency of assistance provided to the user) were collected using a home-based questionnaire. The questionnaire was developed based on previously published studies [13, 16, 17] by our team and based on the feedback gained during preliminary consultations with various stakeholders (c.f., Individual interview). To gain high-level insights into the social impacts of the JACO arm, caregiver participants were also asked to rate 1) the impact of JACO on the caregivers’ perception of user safety when the user is alone, and 2) on the user’s ability to remain in the community rather than being institutionalized. Users were scored on these questions using a 0-10 scale. Caregivers completed the Caregiver Assistive Technology Outcome Measure (CATOM), a reliable assessment tool (intraclass correlation coefficient: 0.86; 95% CI: 0.60–0.95) developed to assess outcomes related assistive technology as perceived by caregivers [18]. The CATOM consists of 1) a series of questions (14 items) investigating the stressors arising from specific caregiving activities or situations, and 2) a series of questions (4 items) assessing the broad impact of caregiving on a caregiver’s participation in life activities and mental health [18]. Caregivers chose two activities for the first series of questions: the activity associated with the greatest caregiving burden and an activity for which they expected the JACO arm to decrease their caregiving burden. For each question, participants had to respond to both a burden scale and a change scale. On the burden scale, participants indicated how frequently they experience each issue (from 5: never to 1: almost always) [18]. For the change scale, they indicated whether each issue had improved, deteriorated, or remained unchanged since the acquisition of JACO, using a 5-point scale (5: improved significantly, 3: no change, 1: deteriorated significantly).

### Data analysis

Descriptive statistics were computed for all quantitative measures (demographic data, CATOM, home-based social impacts questionnaires). All interviews were fully transcribed verbatim. Transcripts were then analyzed using a hybrid deductive-inductive coding process inspired by the approach reported by Fereday & Muir-Cochrane [19]. First, a codebook was developed based on the taxonomy by Jutai et al. [5]. The codebook was pilot tested by one of the authors (JB) with the support of a research associate who is a social worker experienced in qualitative research with people living with a disability. JB is an occupational therapist who had previous experience in qualitative research related to assistive technology use [20]. Next, transcriptions were deductively coded according to the themes within the previously described codebook. Subsequently, data within the social significance domains (Human Assistance, Costs and Service Utilization, and Residential Care Placement), was analyzed using an inductive approach by a fluent French speaker within the research team (MC) (all raw transcripts were in French). Following the initial round of inductive coding, all relevant statements were translated into English by a bilingual (French and English) research personnel (MC) and edited collaboratively with a native English speaker (JF). This translation facilitated for collaborative analysis among both French (MC, JB) and English-speaking (JF) authors. All codes and supporting statements were reviewed by 2-3 authors, and any modifications were discussed until consensus was reached.

Since all participants in this study were recruited from a single region and given the relatively small number of participants in this region, their excerpts were minimally modified (e.g., no indication of the participants’ gender in reported quotes) for confidentiality reasons, in accordance with ethical guidelines. Additionally, for the same reasons, participants are not numbered in this paper to prevent identification.

## Results

### Social significance of JACO outcomes: Qualitative findings

The general topics were described in the codebook as ***Human Assistance*** and ***Cost and Use of Resources***. The ***Human Assistance*** theme encompasses three general nodes: *Favorable Effects, Absence of Effects, and Adverse Effects*. Several topics emerged describing beneficial caregiver outcomes seen with use of JACO, which were then balanced with areas where no impact was perceived or where JACO use created barriers in day-to-day functioning. Alongside variations in perceptions and experiences between participants, intra-participant variation was also observed. ***Cost and Use of Resources*** included several nodes such as *Absence of Effects* (relative to cost and resource use), *Additional Costs, Saving, and Use of Occupational Therapist and Expertise Resources*. Both general topics have been reported in an integrated manner to reflect the interconnected impact of these considerations. Tables 2 and 3 summarize themes for the ***Human Assistance*** and ***Cost and Use of Resources***, respectively.

**Table 2.** Themes associated with *Human Assistance*.

| *Human Assistance* |  |  |
| --- | --- | --- |
| **Favorable effects** | **Absence of effects** | **Adverse effects** |
| Improved sens of security | Activities, life of the caregiver | Complications caused by JACO |
| Changes in the caregiver relationship | Help given despite JACO | The caregiver had to raise funds to get JACO |
| Changes in the help provided | Ability to leave the user alone | Concerns not to break JACO |
| Offer respite | No additional workload or new responsibilities |  |
|  | Relation with others |  |

**Table 3.** Themes associated with *Cost and Use of Resources*.

| *Cost and Use of Resources* |  |  |  |
| --- | --- | --- | --- |
| **Additional costs** | **Absence of effects** | **Savings** | **Use of occupational therapist and Expertise resources** |
|  | No adaptation was necessary to facilitate the use of JACO |  |  |
|  | JACO did not generate any additional costs |  |  |
|  | JACO did not save money |  |  |

### Positive perceptions

#### Human Assistance

*Favorable Effects* included examples where JACO supported quality of life. Caregivers and users discussed their experiences in which the JACO arm impacted the nature, type and amount of assistance provided by the caregiver or another attendant. Specifically, changes were observed in the frequency of caregiver requests and the number of tasks requiring assistance. Caregivers reported experiencing a reduced need to support JACO users, evidenced in statements such as:

User: *“Before, I was always needing to ask, let’s say uh … I’m thirsty, he/she brought me something to drink. It takes me an hour to drink a coffee when someone makes me drink. With JACO, it takes less time. I can drink at my own pace.”*

Caregiver: *“What the JACO brought the most was personal autonomy for the user, but also for the caregiver. Because even if the user needs help with 80-85 % of things, the small 15% is worth 100%.”*

Indeed, the caregiver role in many instances was minimized with the use of JACO.

Caregiver: *“I just have to open the door and drop him/her off there at the cinema, sit back in my car and leave. Like a normal mother/father.”*

The use of JACO was also found to enhance caregiver perceptions of safety regarding the user. With a greater sense of safety, some caregivers appeared to be more comfortable leaving the user alone in the community

Caregiver: *“Before, I was always worried when I left the home, because I didn’t know how he/she would manage himself/herself. Now with JACO, I know that he/she can. If he/she drops something on the ground, JACO will pick it up.”*

Caregiver: *“I can let him him/her go out alone, I don’t worry because I know that, with his/her JACO arm, he/she will be able to open the doors or pick something if it felt on the ground.”*

Caregiver: *“So you know, I am certainly less worried about going to do my things even if I am not with him/her. I know that he/she is capable of managing a lot by him/herself”*.

This idea of improved security was complemented by statements about the JACO user’s ability to open doors without assistance, to manage a mobile phone, and ultimately to live with greater autonomy.

Caregiver: *“The phone was my main concern. If something happens, if he/she can’t access the phone to call me or to call someone else, what will he/she do? With JACO, when I leave, even if something were to happen, I know that he/she can access his/her phone.”*

Another specific area described by users and caregivers was positive change in the caregiver’s relationship to the user. In some instances, the use of JACO enabled family members or significant others to transition from their role as caregivers to a more normalized social role. For example, some parents who are also caregivers shared that the JACO arm allowed them to assume a more typical parental role.

Caregiver: *“I drop him/her to the restaurant with his/her friends and I say, ‘bye-bye and if there is something you can text me.’ Then, I come back home. It’s a more normal relation, like that of a father/mother and their young adult child.”*

Some users also reported that, because of the JACO arm, others within their social circle did not need to carry out caregiver tasks when they were together.

User: *“But it becomes irritating at some point for the boy/girlfriend even if he/she loves you and everything and you love him/her. It creates a long-term divide. Well, JACO means that I don’t need to scrap my relationship. Basically, it gives me autonomy. He/she no longer becomes the attendant.”*

Similarly, JACO was also described as reducing tension in social relationships by improving the quality of time spent with others in social contexts.

*Caregiver: “No well, I spend no less time with him/her. It’s just that the time spent is of better quality because the time I spend with him/her is fun."*

Some caregivers described improved well-being, noting they had more time for respite and to engage in typical, and perhaps personally enjoyable activities.

Caregiver*: “I have time to take my coffee, to do the dishes while he/she is upstairs, doing his/her things. He/she can drink on his/her own. When it’s a smoothie, there is no problem. He/she listens to his/her TV. He/she is on his/her own upstairs. I can do other things.”*

Caregiver: *“The only thing is that it allows him/her to be able to go out of the house, to do something else. So, I can have that time for respite.”*

In summary, the qualitative perspectives on the benefits gained through the use of JACO included greater support to the caregiver role, improved social relationships, a heightened sense of security for the JACO user, and enhanced quality of life for the caregiver.

#### Cost and Use of Resources

It is important to note that few participants reported positive impacts of JACO regarding ***Cost and Use of Resources***. In fact, significant savings due to JACO were reported by only two users. The positive node within ***Cost and Use of Resources*** division (*Savings*) focused on costs of living that were avoided due to JACO use. In one instance, the user’s family chose not to invest in home automation because the user could use the elevator with the help of JACO. This decision was reinforced by the user’s limited life expectancy, reducing the necessity for extensive home automation investments.

In the second case, the user perceived that the autonomy gained with JACO led to significant reductions in external care provision. It is noteworthy that the users who made these statements received their WMRA through a foundation, which naturally influenced the cost-effectiveness of the device in their situation.

Finally, the *Use of Occupational Therapist and Expertise Resources* node encompassed both *Acquisition Process* and *Use of JACO* themes. Statements within these themes focused on the involvement of external resources (i.e., clinicians and vendors) needed to utilize WMRA. There were few positive perceptions related to this node.

### Neutral perceptions

#### Human Assistance

*Absence of Effects* was used to describe examples in which participants clearly stated that the JACO use did not result in meaningful changes on the nature or frequency of human assistance provided by the caregiver or attendant. This theme was therefore descriptively recorded as *Help Given Despite WMRA.* It included situations in which the JACO arm was not utilized in day-to-day activities. In some cases, perhaps due to habit, convenience, or preference, it appears that caregivers provided support rather than relying on the JACO arm.

Caregiver: *“Well, does it help me, personally? Not really. When I am here, I act like the arm of User. If he/she needs something, I give it to him/her.”*

User: *“The doors, I prefer to ask someone, because they’re not automatic. It’s more out of habit. I don’t really open doors with the arm.”*

Other statements indicate more broadly that the JACO arm simply has little impact on the requirement for human support.

Moderator: *“Is JACO changing something on the form or quantity of help you receive?”*

User: *“No, it doesn’t change anything.”*

In some instances, this appears to be because the areas of need lie outside of the JACO’s actual or perceived functional capacity.

User: *“I still need help. I have attendants coming. That doesn’t change because there are things JACO cannot do, like dressing and bathing, those are things that JACO cannot do.”*

In other instances, the user still had the ability to perform daily activities independently within the JACO’s functional capacity. Therefore, caregivers do not assist the user with those activities, even in absence of JACO.

Caregiver: *“There is still a little bit of autonomy, so he/she doesn’t use JACO much. So, if he/she was stuck in his/her wheelchair, we would have to feed him/her, give him/her a drink, etc. And if JACO could do it for us, that would help us. We are not at this stage yet. For the moment he/she can do it alone.”*

Another topic that raised within this category was the lack of effect on the caregiver’s activities and life. In some cases, the caregiver continues to engage in care provision activities:

Caregiver: *“I cannot leave whenever I want. That does not change either. There must be someone close to him/her. We cannot leave him/her alone, but it’s not because … it’s more because of the ventilator. It’s more a question of a risk of ventilator failure”*

#### Cost and Use of Resources

*Absence of Effects* within the ***Costs and Use of Resources*** division continued in a neutral tone. Many users described their experiences with JACO where neither additional savings, nor additional expenses were incurred.

User: *“Well, a finger broke once but we didn’t have to pay anything to fix it.”* Moderator: *“Okay. And were there other events for you to have spent money in connection with JACO?”*

User: *“No”*

In at least one case this was because the JACO arm was acquired as a loan from a rehabilitation organization, and therefore the expenses associated with purchasing and repairing the JACO arm fall outside of the responsibility of the user.

Moderator: *“JACO, did you spend money on it? Aside from the purchase of JACO, did you have to spend money?”*

User: *“It’s not mine.”*

Moderator: *“It’s not yours?”*

User: *“No. It is at the foundation. It belongs to them.”*

In another example, the user made clear that no savings were incurred with the use of the JACO arm.

Moderator: *“Do you think that there are opportunities when it has saved you money? That since you have JACO you didn’t have to buy other stuff, or other things, or that it changed the services you need?”*

User: *“No, no I don’t think so”*

In summary, there were both social and economic perceptions in which the JACO arm neither improved nor worsened the previous status of the user and caregiver.

### Negative perceptions

#### Human Assistance

*Adverse Effects* was used to describe situations in which the JACO arm was perceived to be more of a hindrance to function than a solution. The descriptive theme for this node was *Interference with the Help Provided*. Several caregivers and users described their JACO as a hindrance in various functional areas such as during transfers and vehicle transportation.

User: *“When someone wants to take me, lift me and transfer me, JACO is in the way because it can hit this person.”*

User: *“Tying it up in the truck is a little bit tiring […]. Still, pulling JACO off and putting it back is a bit annoying”*

There were also instances where the WMRA complicated daily activities such as dressing and feeding, either due to its placement or by obstructing caregivers as they assisted the user.

User: *“Yeah. But of course, putting on a sweater is more complicated with the JACO arm than without.”*

Moderator: *“Because it’s like in the way?”*

User: *“Yes that’s it, it’s in the way. We could take it off, it’s an Allen key, you turn it and then you lift your arm, but the arm is about 20 to 25 lbs. It’s not fun for people.”*

Caregiver: *“Yes, to be frank with you, I sometimes hit it when I come to bring the fork to his/her mouth”*

#### Human Assistance x Cost intersection

Other areas of discussion categorized as adverse effects are situated at the intersection of the *Human Assistance* and *Cost* main nodes. Indeed, due to the significant costs associated with the purchase of the JACO arm, several participants discussed extensive efforts over time to raise funds in order to acquire it.

User: *“Well, my dad saw a video on Facebook, and he said my son/daughter needs this. And then my cousin did a fundraising activity, organized a walk. He started from one city and went to another city. He did a part by walking and the other part with his canoe. And my brother did a benefit dinner.”*

Caregiver: *“In two years, I collected $3,500” (note for context, JACO’s cost was approximately $35K-$40K at that time)*.

Caregivers also expressed concerns with breaking or damaging the JACO arm, and how this concern affected its use and function.

Caregiver: *“It’s just that it’s an expensive device, so we must be careful. You don’t want to knock it around. Sometimes, when there are a lot of children, we will put it away if User isn’t using it. But it’s rather rare.”*

Some caregivers were anticipating some challenges and anxieties regarding the end of life of the JACO arm. Some of these concerns revealed a tension between their desire to provide the best opportunities for the user and the important efforts required to raise funds for purchasing a new JACO.

Caregiver: *“As JACO is now 7 years old, the doctor told User that we should start looking for fundraising campaigns if he wants a new one. But I am tired, and I told ‘Ah no, it’s not true!’. But then User talked to me again about it [later]. He wants a new one for sure.”*

Another caregiver experienced an incident due to the failure of one of JACO’s motors reaching its end of life. This raised moderate safety concerns for the user that were not present before the incident.

Caregiver: *“Now [since the incident], I call someone close by when I leave, to say: ‘Can User call you if JACO does not work?’ Otherwise, when everything is ok, I do not need to do that. He/she has his/her JACO arm’s joysticks, everything is fine, he/she is able to go out, able to call, he/she is able to do all his/her things.”*

#### Costs and Use of Resources

The ***Costs and Use of Resources*** node, *Additional Costs*, focused on instances where the use of JACO incurred additional expenses for the user, caregiver, or society. Examples included the need for modifications or adaptations. In our sample, this was mostly homemade modifications of manipulated objects, such as home-sewn straps to make opening doors easier.

User: *“Like the door handle of my fridge, I put a two feet long tie-wrap around it and fixed it with electric tape.”*

JACO users also incurred additional costs through repairs or maintenance. Some also anticipated the costs that will be required at the JACO arm end of life.

User: *“You have the maintenance, the cost and then you have repairs for an arm that will not last more than five years. We apologize, but we hadn’t realized that the engines would not be as good. So, there you’d be better off having a second generation.”*

Some costs associated with the use of a JACO arm appear to have positive implications. For example, the purchase of a smartphone became necessary once the WMRA enabled the user to manage a phone more autonomously.

### Quantitative Findings

JACO users’ mean score in response to question of whether the JACO arm affected caregiver perception of user safety was moderate: 5.5/10 [SD= 3.8]. Caregivers’ mean score was 6.1/10 [SD=3.6]. However, most disagreed that the JACO arm contributed significantly to their ability to remain in the community rather than being institutionalized (users’ mean score: 2.9/10 [SD=3.1]; caregivers’ mean score: 4.3/10 [SD=3.3]). As shown in Figure 1, most caregivers reported little to no positive impact (Change scale: 3 ≤ CATOM < 4) of the JACO arm on the burden associated with aiding across both targeted activity items and global assessment items of the CATOM. On average, the participants reported “*Sometimes*” for each selected activity related to burden (Difficult activity: 3.1 [SD=0.4]; Activity with expectation: (3.4 [SD=0.7]) and globally (3.2 [SD=1.1]), although there were large variations within the sample.

**Figure 1.**
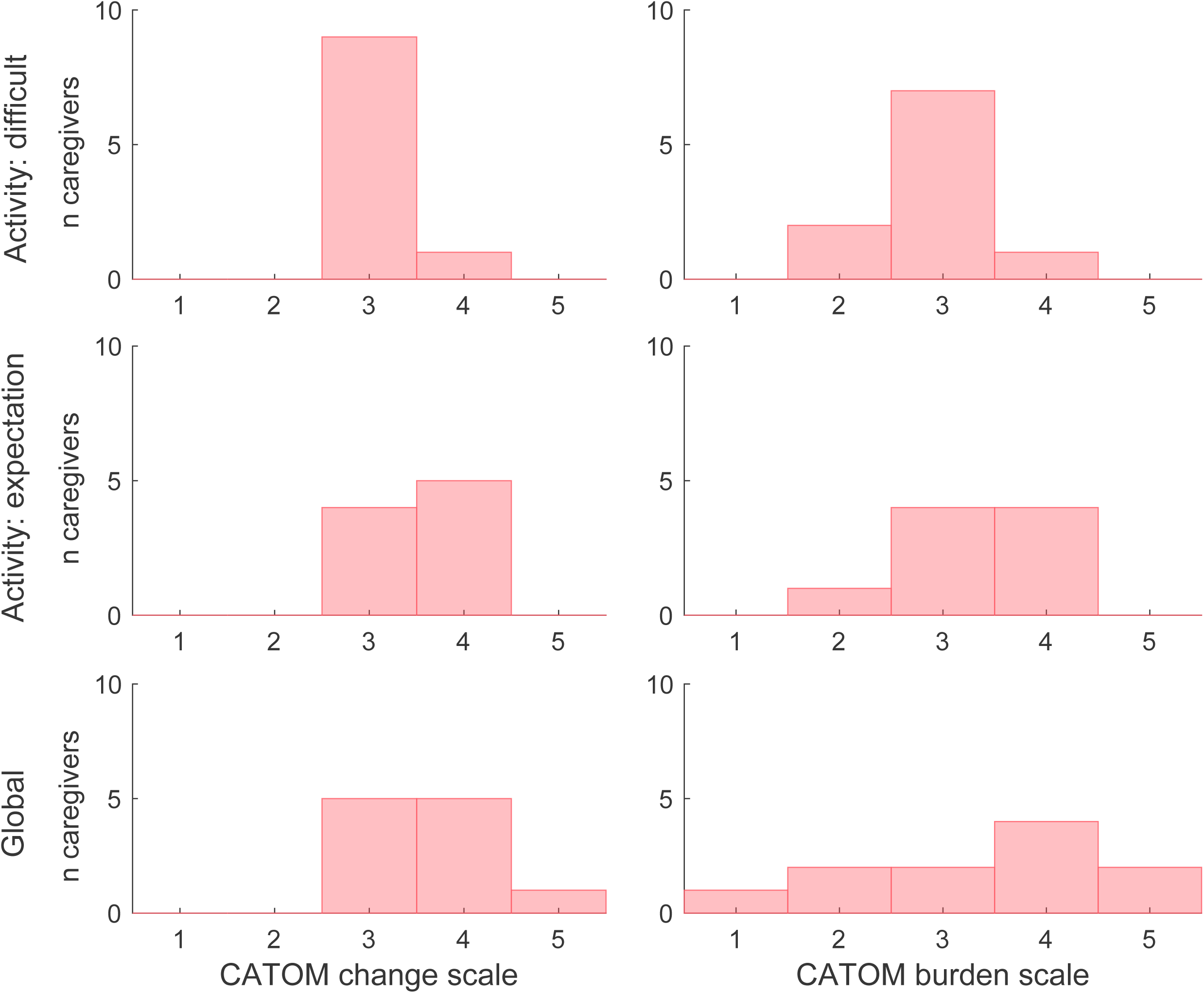

## Discussion

### Technology implementation

The present study assessed the social impact of JACO as perceived by a sample of users and caregivers. Differences in satisfaction with JACO use, both among users and across activities, highlighted the importance of approaching each user as an individual with unique needs, contexts, experiences, and preferences.

The HAAT model emphasizes the importance of considering not only the technology and the user, but also the activities in which they engage, and the physical, social, and cultural context in which the user lives their day-to-day life [6]. Our findings align with a holistic perspective of assistive technology, emphasizing the impact of activity, social support, and physical context.

Therefore, careful attention should be paid to the anticipated user’s activity-related goals, as well as their home and community context, when deciding whether a WMRA will have a significant impact on the user’s day-to-day life.

Furthermore, barriers in the physical environment (such as vehicle measurements, surface heights, and lift parameters) should be identified and addressed, if possible, as part of the assistive technology service delivery process [21].

### Perceptions illuminating social impact

Both user and caregiver feedback indicated that our sample perceived the JACO arm to have a moderate and variable impact on safety when the user is alone. JACO can be used to access and operate different devices critical to one’s safety, such as picking up a phone or opening a door. Inability to manage such things can render an individual highly vulnerable in emergency situations. However, participant feedback indicates that the capabilities facilitated by the JACO arm were not always sufficient to overcome every barrier to safety. Therefore, it is important to access the context of each individual user in relation to the functionalities that the WMRA enables. JACO was reported to enhance satisfaction in many aspects of both users’ and caregivers’ quality of life. Increased autonomy and perceptions of safety were complemented by redefined relationships between caregivers and care recipients. This underscores both the impact of caregiving tasks on self-identified roles and the potential downstream social impact of improved ability to function autonomously in daily life. The ability to switch from the role of caregiver to that of mother, father, or significant other can be highly meaningful to both the caregiver and user. Furthermore, the relational impact of a WMRA emphasizes the importance of considering both the user and the caregiver in WMRA service delivery.

The perceived impact of the JACO arm on users’ ability to remain in the community setting was rather low, indicating that the JACO arm alone was not a determinate of day-to-day life context. This finding seems to be related to, not a lack of functionality, but the presence of alternative compensatory strategies (for example, the support of paid and unpaid caregivers). In fact, some participant feedback indicated that the assistance that users receive from caregivers could not be replaced with a WMRA. This is consistent with previous research by Beaudoin et al. [16], which reported that using WMRA for personal care activities was met with either “some” or “a lot” of difficulty and that most users reported that personal care tasks were completed via human assistance rather that with the WMRA [16]. Within the present study, in some cases, previous habits or the burden associated with the use of JACO seemed to influence the choice of caregiver assistance versus JACO assistance. This finding is not inherently negative but rather informative about situations where the convenience or autonomy of WMRA use does not surpass that of human assistance. In fact, referring back to the previously mentioned investigation by Beaudoin et al. [16], several users who reported difficulty with JACO in personal care tasks also expressed satisfaction with the device, indicating that complete independence in using JACO was not necessary for it to be perceived positively [16]. By better understanding which activities are effectively supported by WMRA versus those better addressed by human assistance, clinicians will be better equipped to determine the appropriateness of WMRA for new potential users based on their habits, needs, support systems, and goals. The addition of a WMRA can contribute to increased autonomy. However, healthcare providers assisting in the acquisition and training of the WMRA should discuss realistic expectations regarding independence or potential changes in living contexts. This allows users and caregivers to anticipate the functional abilities and social factors that will likely be affected, as well as those that may not. Of note, several participants reported limited involvement of occupational therapists in the JACO acquisition process.

While this approach resulted in lower costs, it may also have led to suboptimal use of the JACO arm or, more worryingly, the selection of an inappropriate assistive device for the individual client’s needs.

Furthermore, since there is limited evidence of cost savings among JACO users in our sample, careful consideration should be given to the cost-benefit analysis of recommending expensive devices like JACO. It is important to incorporate key clinical stakeholders into the WMRA service delivery process to conduct further research aimed at identifying and recommending effective and financially prudent guidelines for WMRA implementation. Determining user satisfaction with assistive devices is of paramount importance to their adoption and sustained use. Even well-designed assistive devices lose their effectiveness if they are not utilized.

Incorporating user perspectives into the ongoing refinement of assistive devices can ultimately enhance the value of care delivered to device users [22, 23].

This study provides novel insights into a growing body of evidence on stakeholder perceptions of using JACO. While previous studies have analyzed the potential value and use of WMRAs in small samples [11, 12], the present study scrutinized the perceived social impact of WMRA use among several users and caregivers. Investigations into the perceived effectiveness of assistive devices can provide valuable insights, beyond clinic-based objective measures, shedding light on how the device is utilized outside of clinical settings.

Understanding instances where the JACO arm is perceived as a barrier to function is crucial. In many cases this was related to logistical challenges associated with adding a sizable item to the user’s wheelchair. The added size and weight of the JACO arm, beyond the structure of the wheelchair, should be considered in relation to the user’s home environment (e.g., door width, shelf height, etc.) and in terms of the ongoing assistance caregivers will provide. For instance, new JACO users and their caregivers may benefit from additional training on completing a variety of transfers once the JACO arm is integrated into the wheelchair. Some users may prefer removing the arm prior to transfers, while others may be able to position the arm in such a way that will not impede the transfer. In either case, additional attention to preparing the user and caregiver for anticipated and potential logistical challenges associated with WMRA use may improve overall satisfaction with the device.

## Limitations

This study has several limitations that need to be disclosed. First, the observational mixed-method research design was exploratory and was chosen to gain insight into both the breadth and depth of the impact of a WMRA within the studied population. It provides an overview of the perceived social significance of JACO outcomes but does not allow for a definitive interpretation of the relative impact of this assistive device compared to other solutions. Second, evaluator subjectivity and the risk of response bias by the participants are inherent to qualitative interview methods.

The first limitation was addressed through a multi-level consensus process of interpretation involving three separate researchers who reviewed and discussed transcripts, ultimately reaching consensus on the data interpretation. Furthermore, the fact that participants reported both positive and negative impacts of the JACO arm supports the validity of the obtained data. The generalizability of this study is limited, considering that our participant sample is exclusively from Canada. Therefore, participant perspectives may have been affected by region-specific reimbursement policies. In addition, reasons for non-participation were not investigated. Thus, perspectives of individuals with negative opinions, or who discontinued use of their JACO may have been overlooked. Future studies should continue to investigate additional sample populations to expand the research on the use of JACO and evaluate potential cofounding factors such as whether the duration since acquiring a JACO arm affects its impact on the daily lives of users and caregivers.

## Conclusion

The present study assessed users’ and caregivers’ perspectives on the outcomes of the JACO arm in social significance domains using both quantitative assessment and in-depth qualitative interviews in order to attain a holistic picture. Qualitative interviews covered two areas of social impact, 1) ***Human Assistance***, which investigated JACO’s impact on the caregiver’s daily life and quality of life, and 2) ***Cost and Use of Resources***, which considered the financial impact, both positive and negative, associated with JACO use. Findings indicated that experiences vary both among and within caregivers.

## Data Availability

All data produced in the present study are available upon reasonable request to the authors

## Acknowledgements

We would like to thank David Pacciolla, Anissa Bastien, Daphnée Dumouchelle, Valérie Filion, Marie Gagnon, Marika Girard, and Josiane Lettre for their support at various stages of the study. We would also like to express our gratitude to all participants who took part in this study and in preliminary consultations.

