## Supplementary material 1 for "Social impact of the JACO wheelchair-mounted robotic arm on users and their caregivers"

**Main questions in the interview guide for the large study in JACO arm users**

1. Why did you buy a JACO arm?

2. What activities do you perform with the JACO arm?

3. In what contexts do you use the JACO arm? For example, do you use it only at home, or also at work and in public places?

4. How has your ability to use the JACO arm evolved since you acquired it?

5. What are the impacts or effects of the JACO arm on your caregivers?

6. How does the JACO arm influence your relationships with others?

7. How does the JACO arm influence your sense of well-being?

8. Are there any advantages of the JACO robotic arm that you experience but we haven't discussed?

9. What disadvantages do you experience with the JACO arm?

10. Has using the JACO arm led to personal expenses or savings for you?

11. Have there been periods when you used the JACO arm more or less than usual?

12. What factors influence your use of the JACO arm and its impact?

13. What technical features would make it easier for you to use the JACO arm?

14. What support would you like to have for using the JACO arm that you haven't received?

15. If your JACO arm were taken away from you indefinitely, how would you react?

16. If you were offered to exchange the JACO arm for the services of a paid caregiver who would be permanently available, how would you react?

17. What advice would you give to someone who has recently acquired a JACO robotic arm?

18. What criteria do you think should determine whether the JACO robotic arm is assigned to the right person?
