## Supplementary material 2 for "Social impact of the JACO wheelchair-mounted robotic arm on users and their caregivers"

**Main questions in the interview guide for caregivers regarding the large study on using the JACO arm**

1. What were your expectations or concerns when your loved one acquired JACO?

2. In your opinion, what impact does JACO have on the life of your loved one?

3. How has the impact of JACO evolved since they acquired it?

4. What activities does your loved one perform with the JACO arm?

5. How does JACO influence their sense of well-being?

6. What disadvantages does your loved one experience with JACO?

7. How has JACO changed or affected your personal life?

8. How do you feel about the safety of your loved one since they acquired JACO?

9. How does JACO influence the assistance you provide to your loved one?

10. Are there situations where JACO either complicates or facilitates your assistance to your loved one?

11. What impacts or effects does JACO have on your daily activities and lifestyle?

12. How does JACO influence your own sense of well-being?

13. How does JACO influence your relationships with your loved one?

14. What disadvantages do you experience personally due to JACO?

15. Has JACO led to any personal expenses or savings for you?

16. Are there any advantages or disadvantages of your loved one's JACO robotic arm that we haven't discussed?

17. What support related to JACO would you have liked to receive but have not received yet?

18. If your loved one's JACO arm were taken away indefinitely, how would you react?

19. What advice would you give to a caregiver of someone who has recently acquired a JACO robotic arm?
